# Change in gut microbiome precedes symptomatic Alzheimer’s disease and is correlated with amyloid-beta accumulation in the brain

**DOI:** 10.64898/2026.10.15.26365677

**Authors:** Andrew P. Shoubridge, Sarah K. Manning, Sophia Tan, Sarita Kidson, Alyson Richard, Julie Trinder, Sophie Miller, Kate Laver, Miia Rahja, Joanne Robertson, Maria Crotty, Steven L. Taylor, Geraint B. Rogers

**Affiliations:** Microbiome and Host Health Programme, South Australian Health and Medical Research Institute (SAHMRI), Adelaide, South Australia, Australia; College of Medicine and Public Health, Flinders Health and Medical Research Institute, Flinders University, Bedford Park, South Australia, Australia; SAHMRI, Adelaide, South Australia, Australia; Central Adelaide Local Health Network, SA Health, Adelaide, South Australia, Australia; Australian e-Health Research Centre, Commonwealth Scientific and Industrial Research Organisation (CSIRO), Queensland, Australia; College of Nursing and Health Sciences, Flinders University, Bedford Park, South Australia, Australia; Southern Adelaide Local Health Network, SA Health, Adelaide, South Australia, Australia; Registry of Senior Australians Research Centre, SAHMRI, Adelaide, South Australia, Australia; The Florey Institute, The University of Melbourne, Parkville, Victoria, Australia

**Author notes:** Correspondence: Professor Geraint Rogers, South Australian Health and Medical Research Institute, North, Terrace, Adelaide, SA 5001. Australia.

**Keywords:** Alzheimer’s disease, amyloid-P, gut microbiome, preclinical, presymptomatic

## Abstract

**INTRODUCTION:** Alzheimer’s disease (AD) begins with cerebral amyloid-beta (AP) deposition decades before clinical symptoms. While the microbiota–gut–brain axis has been implicated in AD, its role in this preclinical phase remains unclear. We examined potential links between gut microbiome features and presymptomatic cerebral Ap burden.

**METHODS:** Asymptomatic individuals (n=101, median age 70.0 years, 53% female) underwent cognitive assessment, cerebral Aβ quantification, and stool metagenomic sequencing. Associations between microbial characteristics and Aβ levels were performed using multivariable-adjusted ordinal logistic regression.

**RESULTS:** Gut microbiota composition varied with Aβ levels (p(perm)=0.03, R_2_=0.014). Higher Ap was associated with increased *Alistipes communis,* and decreased *Anaerotignum faecicola* and *Eubacterium rectale,* abundance (q<0.001). Aβ-associated microbiome traits corresponded to lower biosynthetic capacity for γ-aminobutyric acid, and systemic regulators of immunity and autophagy (q<0.01).

**DISCUSSION:** Early Aβ accumulation is associated with distinct gut microbial signatures and reduced neuroactive metabolic potential, identifying targets for potential preclinical AD intervention.

## 1 BACKGROUND

Alzheimer’s disease (AD) is the most common form of dementia, accounting for 50-70% of cases worldwide^1^. AD develops slowly, with progression from presymptomatic disease to death typically spanning several decades^2^. The pathological hallmarks of AD, and the basis for its diagnosis, are the accumulation of amyloid-beta (Aβ) plaques and/or tau neurofibrillary tangles in the brain^3^. Current pathological evidence positions Ap deposition as the earliest detectable abnormality, preceding downstream pathological changes and the emergence of clinical symptoms^4^.

Clinically, AD is characterised by a gradual decline in memory and broader cognitive functions, including language, visuospatial skills, and executive function^5^. AD-related pathology in the absence of clinical symptoms is defined as preclinical AD^6^. This early stage of disease development represents a potential window for intervention prior to substantial neurodegeneration.

One potentially important influence on the early stages of AD development is the microbiota–gut–brain axis; the bidirectional communication network linking the gut microbiota, gastrointestinal tract, and central nervous system^7^. Microbiota–gut–brain pathways capable of influencing neurophysiology and cognition primarily include microbial synthesis of neurotransmitters, such as γ-aminobutyric acid (GABA) and glutamate^8^, modulation of systemic immune function through the production of short-chain fatty acids (SCFAs)^9^, and the metabolism of essential amino acids, including leucine and tryptophan^10^. These microbial traits have been associated with dementia severity in later life^11^ and may represent modifiable contributors to dementia risk, including AD, during its earliest stages.

Although numerous studies have identified associations between the gut microbiome and symptomatic AD^12–14^, little is known about the potential influence of host–microbiome interactions during the preclinical phase. Our aim was to relate the gut microbiome to Ap accumulation in the brains of individuals with preclinical AD, with a specific focus on microbiome taxonomic and metabolic traits that can influence host immunity and neurophysiology.

## 2 METHODS

### 2.1 Study design and cohort

Study participants were recruited from the Australian Dementia Network (ADNeT) Trials Ready Cohort (TRC) through the Screening for Trials Initiative^15^. ADNeT enrols participants aged 40 years and older with a Mini-Mental State Examination (MMSE) score >15. All participants who consented for this study underwent neuropsychological assessment to measure cognitive impairment, positron emission tomography (PET) imaging to measure cerebral Ap burden, blood collection for circulating AD marker levels, and stool sample collection.

### 2.2 Clinical data collection and definitions

Anonymised participant data, including the demographic variables, Aβ values, and AD classifications, were collected from the ADNeT TRC^15^. Ap accumulation was determined by PET imaging. Aβ quantification utilised the Centiloid (CL) scale in which levels correspond with histopathologic classification and correlate with prognosis^16^. In this scale, “0” corresponds to the mean scan measure of healthy young adults without Aβ deposition, and “100” corresponds to the mean scan measure of individuals with mild AD. A score of “25” is the commonly employed threshold for AD pathology^16^.

### 2.3 Metagenomic analysis of stool

Stool samples were collected in Norgen Stool Nucleic Acid Collection and Preservation Tubes (Norgen Biotek, Thorold, ON, Canada). Microbial DNA was extracted using the PowerLyzer PowerSoil DNA Isolation Kit (Qiagen, Hilden, Germany) as previously described^17^.

Indexed, paired-end DNA libraries were prepared using the xGen DNA Library Prep Kit (Integrated DNA Technologies, Singapore) and sequenced at a depth of 6 Gb on an MGI G400 platform with 150 bp paired-end reads. Forward and reverse sequences were quality-filtered using Trimmomatic (v0.39)^18^, and human reads were removed with Bowtie (v2.5.3) against the NCBI human reference genome release GRCh38^19^. Taxonomic relative abundance was assigned using MetaPhlAn (v4.0.6)^20^, while microbial metabolic pathway abundance was assigned using HUMAnN (v3.9) against the MetaCyc database^21^. Sequence data have been entered into the European Nucleotide Archive (ENA) at EMBL-EBI under accession number PRJEB109799.

### 2.4 Microbiome characterisation

Diversity metrics were generated from species-level taxonomic relative abundances. Alpha diversity measures included Shannon-Wiener diversity (*H’*), Simpson’s index (*D*), Simpson’s Dominance (Inverse) index (1/*D*), and species richness (*d*), and were determined using the ‘vegan’ R package (version v2.7-1)^22^. Beta diversity analysis was performed using permutational multivariate analyses of variance (PERMANOVA) on Bray-Curtis dissimilarity distances in an unrestricted permutation of unfiltered, square-root transformed relative abundance data, by the function ‘adonis2’ from the vegan R package. Principal Coordinate Analysis (PCoA) plots for beta diversity were generated using the ‘vegan’ R package and visualised using “ggplot2” (v.4.0.0).

The functional capacity of the gut microbiota was defined by the genetically-encoded functional traits detected within the metagenome. MetaCyc pathways from HUMAnN that were present in <30% of participants were excluded. Pathways were categorised according to classes mapped within the MetaCyc database as fermentation to short-chain fatty acids, amino acid biosynthesis, neurotransmitter biosynthesis, or other.

### 2.5 Statistical analysis

Statistical analysis of participant demographics was performed using the Aβ CL value as a categorical variable (classification of AD for Ap >25 CL) utilising two-tailed Mann-Whitney tests for continuous variables and Pearson’s chi-square tests for categorical variables.

Both unadjusted and multivariable regression models were employed in all other analyses, using the Aβ CL value as a continuous variable. Multivariable-adjusted models accounted for age, sex (male or female), education (years), hypertension (history or current), diabetes (history or current), smoking (history or current), obesity (current), traumatic brain injury (history), depression (history or current), high cholesterol (history or current).

Within-individual microbiome variables included alpha diversity metrics, species-level relative abundance (detected in >30% of participants), and metabolic pathway abundance (detected in >30% of participants). All within-individual variables were ranked into tertiles. Ordinal logistic regression was performed to assess the effect of Aβ on microbiome variables using the “MASS” function in R. Odds ratios (ORs) for regression model coefficients were tested for statistical significance *(p* <0.05), using the Aβ CL value as a continuous value for the predictor variable. False discovery rate (FDR) multiple hypothesis testing was conducted with the Benjamini and Hochberg method across all profiles using the “p.adjust” function in R, at a significant threshold of 0.05. Correlations between microbial functional characteristics and Aβ CL values were calculated by Spearman partial correlations (with multivariable adjustment) using the ‘ppcor’ package in R^23^ and tested for statistical significance (*p*<0.05).

## 3 RESULTS

### 3.1 Cohort characteristics

The study cohort included 101 participants (**Table 1**), of whom 53% were female and 38% (n=38) had Aβ levels indicative of AD pathology (Ap >25 CL). Those with AD pathology had a median CL score of 78.1 (interquartile range [IQR] = 42.8, 109.9), compared to-0.7 (IQR =-4.0, 4.6) in those without AD. Those with AD were significantly older (median age AD = 72.0 [67.3, 74.7]; non-AD = 68.0 [62.0, 72.5], *p*<0.01) and less likely to be obese (AD = 11%; non-AD = 32%, *p*<0.05). Cognitive function (MMSE score), years of education, hypertension, diabetes, smoking, traumatic brain injury, depression, and high cholesterol did not differ significantly by AD classification (*p*>0.05).

**Table 1:** Cohort characteristics by level of accumulated amyloid-beta.

|  | **Total**<br>**100%**<br>**(n=101)** | **Aβ <25 CL**<br>**62%**<br>**(n=63)** | **Aβ ≥25 CL**<br>**38%**<br>**(n=38)** | **P-value** |
| --- | --- | --- | --- | --- |
| **Amyloid-β CL Value:**<br>median (IQR) | 6.3 (-1.5, 52.6) | -0.7 (-4.0, 4.6) | 78.1 (42.8, 109.9) | **<0.0001** |
| **Age:**<br>median (IQR) (years) | 70.0 (63.0, 74.0) | 68.0 (62.0, 72.5) | 72.0 (67.3, 74.7) | **<0.01** |
| **Sex:** % (n) |  |  |  |  |
| Female | 52.5 (53) | 47.6 (30) | 60.5 (23) | ns |
| Male | 47.5 (48) | 52.4 (33) | 39.5 (15) |  |
| **MMSE Score:**<br>median (IQR) | 29.0 (28.0, 30.0) | 29.0 (28.5, 30.0) | 29.0 (27.3, 29.7) | ns |
| **Education:**<br>median (IQR) (years) | 12.0 (11.0, 15.0) | 12.0 (11.0, 15.0) | 12.0 (10.0, 14.8) | ns |
| **Hypertension:** % (n)* | 32.7 (33) | 33.3 (21) | 31.6 (12) | ns |
| **Diabetes:** % (n)* | 5.9 (6) | 6.3 (4) | 5.3 (2) | ns |
| **Smoking:** % (n)* | 37.6 (38) | 38.1 (24) | 36.8 (14) | ns |
| **Obesity:** % (n)<sup>#</sup> | 23.8 (24) | 31.7 (20) | 10.5 (4) | **<0.05** |
| **TBI:** % (n)<sup>^</sup> | 6.9 (7) | 6.3 (4) | 7.9 (3) | ns |
| **Depression:** % (n)* | 16.8 (17) | 14.3 (9) | 21.1 (8) | ns |
| **High cholesterol:** % (n)* | 47.5 (48) | 44.4 (28) | 52.6 (20) | ns |

Abbreviations: Aβ = amyloid-beta; CL = Centiloid; IQR = interquartile range; MMSE = Mini-Mental State Examination; ns = not significant; TBI = traumatic brain injury.
\*Current or recorded history of medical condition.
<sup>^</sup>Recorded history only of medical condition.
<sup>#</sup>Medical condition current as at time of enrolment.

### 3.2 Gut microbiota characteristics differ with accumulated amyloid-beta levels

A total of 844 bacterial species, representing 283 genera, were identified in the cohort. Of these, 214 species were detected in >30% of participants. Obesity (Pseudo-F=1.70, R^2^=0.017, *p* (perm)=0.007) and Aβ (Pseudo-F=1.44, R^2^=0.014, *p* (perm)=0.031) explained the greatest portion of gut microbiome compositional variance (**Fig. 1A, B**). No significant associations were found between alpha diversity measures and Aβ accumulation (**Table S1**).

**Figure 1:**
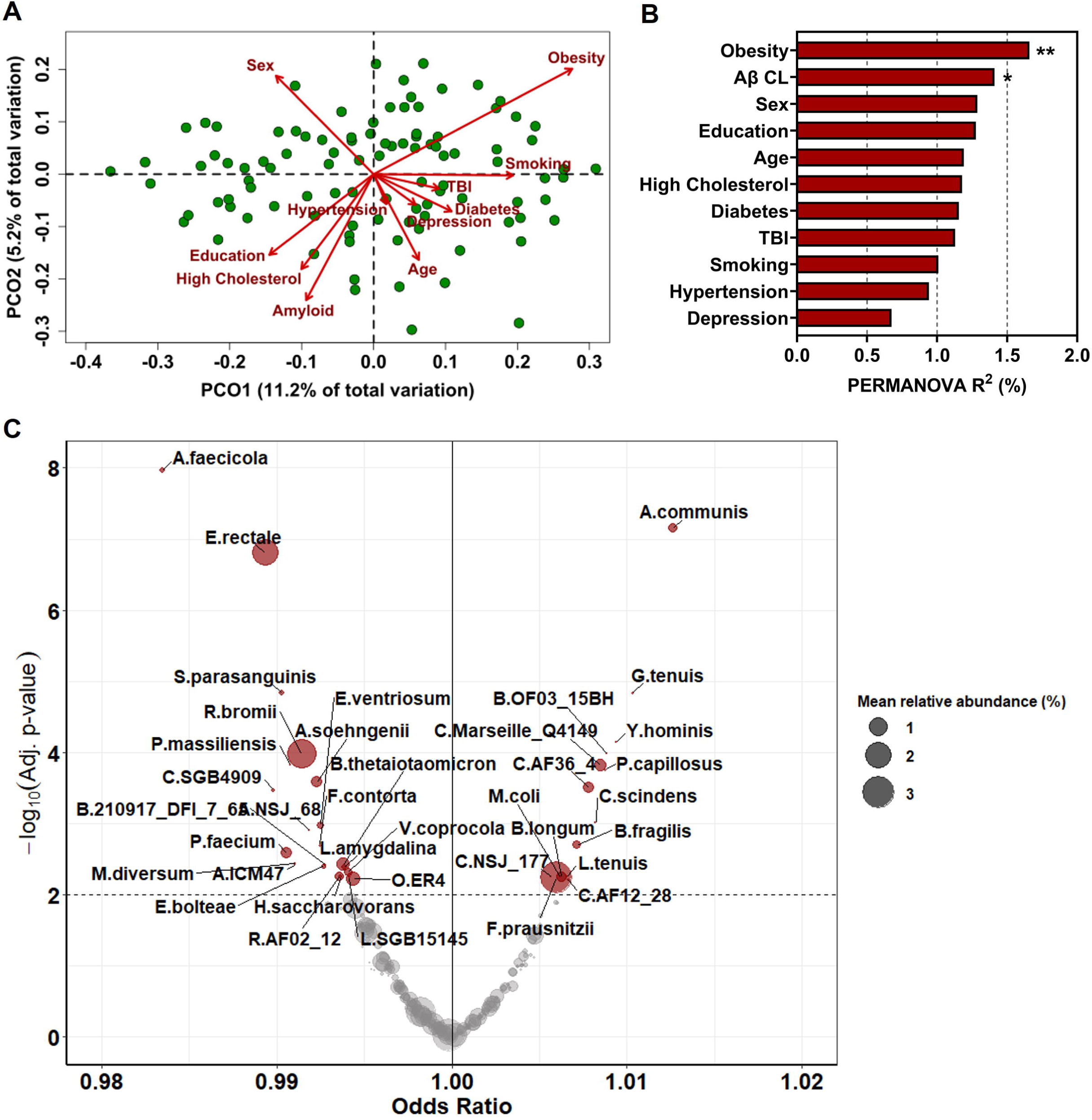
**Species-level differences in the gut microbiome of people undergoing testing for preclinical Alzheimer’s disease (AD) by level of accumulated amyloid-beta (AP). A**) Principal Coordinate Analysis plot of Bray-Curtis similarity matrix for all individuals (*n*=101), with contributing features overlaid and length of arrow indicating the amount of importance. **B**) Permutational ANOVA for Bray-Curtis similarity (beta-diversity) listing features and their significance to variation in composition (R^2^). \**q*<0.05; \*\**q*<0.01. **C**) The mean relative abundance of microbial species related to AP accumulation, presented as odds ratios. Performed by multivariable analysis treating AP as a continuous variable and adjusting for age, sex, education, hypertension, diabetes, smoking, obesity, traumatic brain injury, depression, and high cholesterol. Labelled species were statistically significant (*q*<0.01) following FDR correction.

Of all microbial species identified across the cohort, the relative abundance of 62 species was significantly associated with Aβ (**Table S2**). The strongest negative associations with Aβ accumulation were with *Anaerotignum faecicola* (OR: 0.983 [0.978, 0.988]), *Eubacterium rectale* (OR: 0.989 [0.986, 0.993]), *Ruminococcus bromii* (OR: 0.991 [0.988, 0.995]), and *Anaerobutyricum soehngenii* (OR: 0.992 [0.989, 0.996] (all *q*<0.001; **Fig. 1C**). Amongst the strongest positive associations were *Alistipes communis* (Odds Ratio [OR]: 1.013 [1.009, 1.017]), *Guopingia tenuis* (OR: 1.010 [1.006, 1.014]), *Bacteroides fragilis* (OR: 1.007 [1.003, 1.011]), and *Faecalibacterium prausnitzii* (OR: 1.006 [1.002, 1.010]) (all *q*<0.01; **Fig. 1C**).

### 3.3 Gut microbiome functional capacity varies with accumulated amyloid-beta

Three hundred and eighty-two MetaCyc pathways were detected in >30% of participants, where 26 related to SCFA production, 49 related to amino acid production, and 13 to neurotransmitter production. Of these, 108 were significantly related to the degree of AP burden following adjustment and FDR correction (**Table S3**).

AP levels were significantly and inversely associated with microbial pathways related to the production of SCFAs (*n*=17 pathways), including acetate (FERMENTATION-PWY; OR: 0.992 [0.988, 0.996], *q*<0.001), propionate (PWY-5494; OR: 0.993 [0.988, 0.997], *q*<0.01), and butyrate (CENTFERM-PWY; OR: 0.995 [0.991, 0.999, *q*<0.05]) (**Fig. 2A**). Similar relationships were observed in relation to autophagy-regulating functional amino acids (*n*=19 pathways) including methionine (HSERMETANA-PWY; OR: 0.991 [0.987, 0.995], *q*<0.001), arginine (ARG-POLYAMINE-SYN; OR: 0.995 [0.991, 0.998], *q*< 0.05) and isoleucine (PWY-3001; OR: 0.994 [0.990, 0.997] *q*<0.01) (**Fig. 2B**), and neurotransmitters and neurotransmitter precursors (*n*=4 pathways), including GABA (GLUDEG-I-PWY; OR: 0.995 [0.991, 0.998], *q*<0.05) and tryptophan (PWY-6629; OR: 0.992 [0.988, 0.996], *q*<0.001) (**Fig. 2C**).

**Figure 2:**
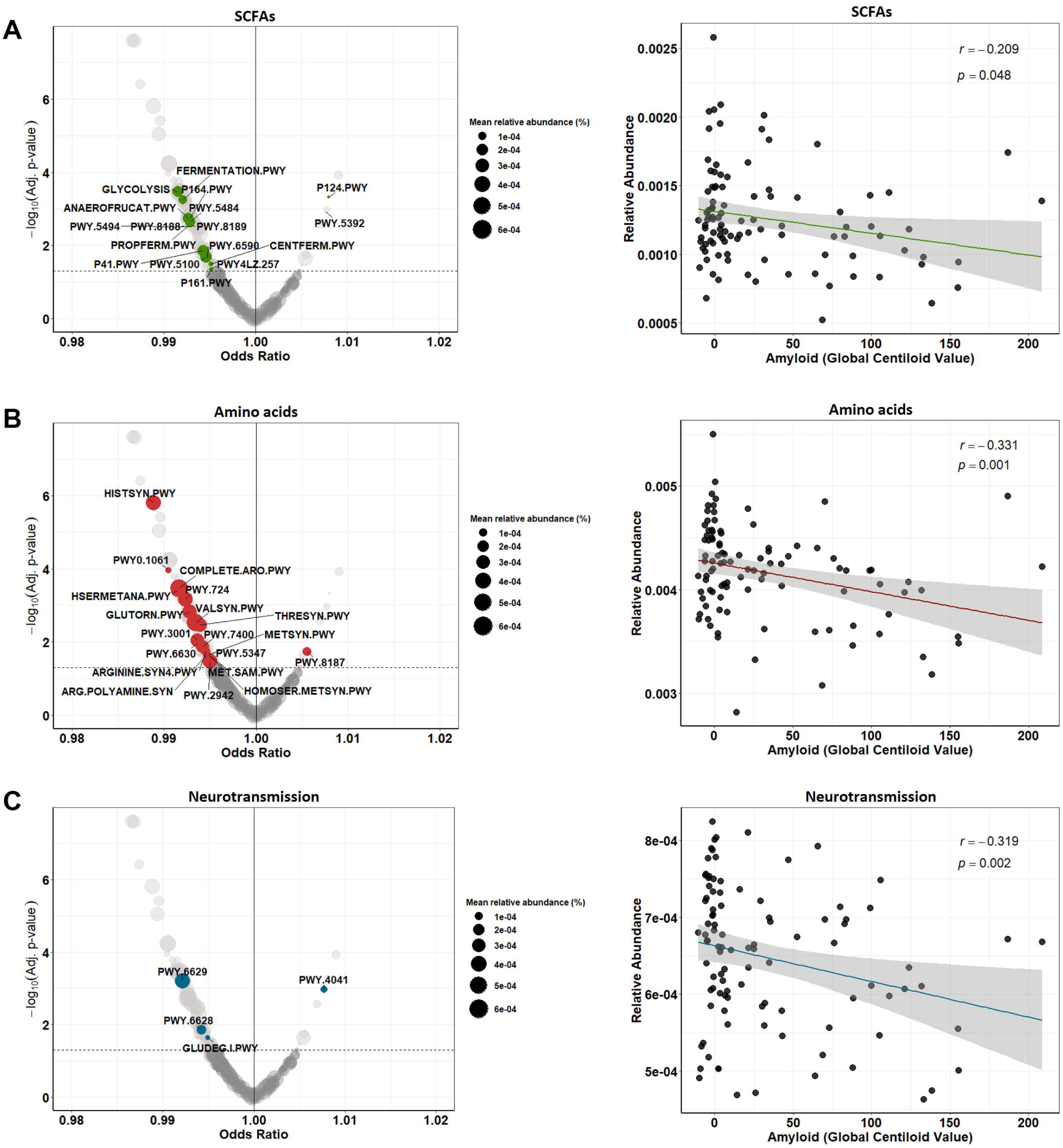
**Functional differences in the gut microbiome of people undergoing testing for preclinical Alzheimer’s disease (AD) by level of accumulated amyloid-beta (AP).** Odds ratios and partial correlations of associations between AP accumulation and the microbial functional capacity to produce SCFAs (**A**), amino acids (**B**), and neurotransmitters (**C**) for all individuals (*n*=101). Performed by multivariable analysis treating Aβ as a continuous variable and adjusting for age, sex, education, hypertension, diabetes, smoking, obesity, traumatic brain injury, depression, and high cholesterol. Labelled functional pathways were statistically significant (*q*<0.05) following FDR correction. Partial regression plots report the adjusted association between the summed relative abundance of the labelled functional pathways and AP accumulation.

A further 68 metabolic pathways significantly differed by level of Aβ, of which 62 pathways were inversely related and 6 pathways positively related to Aβ, with the strongest associations including those related to stachyose degradation (PWY-6527; OR: 0.987 [0.982, 0.991, *q*<0.0001], D-galactose degradation (PWY-6317; OR: 0.987 [0.983, 0.991], *q*<0.0001), and nicotinamide recycling (PWY-7761; OR: 1.009 [1.005, 1.013], *q*<0.0001) (**Table S3**).

## 4 DISCUSSION

In this study, we characterised the gut microbiome of individuals without cognitive impairment who were undergoing assessment for preclinical AD. We identified significant associations between microbiome composition and cerebral Aβ burden. These compositional changes were accompanied by a reduction in predicted microbial capacity to produce immunomodulatory SCFAs, amino acids involved in autophagy regulation, and neurotransmitters and their precursors. While not evidence of causality, our findings add to the growing body of literature implicating the gut microbiome in AD development^12–14^, presenting opportunities for intervention.

We provide evidence that measurable alterations in the gut microbiome are present prior to cognitive impairment, with several bacterial taxa associated significantly with Aβ burden. Notably, Aβ levels were inversely related to taxa widely considered beneficial to intestinal and immune health. These included *Ruminococcus bromii*, the primary degrader of dietary resistant starch^24^; *Eubacterium rectale*, a key contributor to intestinal mucus barrier integrity^25^; *Anaerotignum faecicola*, a degrader of dietary fibre^26^; and *Anaerobutyricum soehngenii*, a metaboliser of bile acids^27^. These taxa are prominent SCFA producers and have been implicated in protection against intestinal inflammation^25^, bacterial infection^26^, and type 2 diabetes^27^. Their depletion with increasing Aβ is consistent with a loss of microbiome-mediated protective functions during the preclinical phase of AD.

In contrast, higher Aβ burden was positively associated with enrichment of *Alistipes communis*, *Guopingia tenuis*, *Bacteroides fragilis*, and *Faecalibacterium prausnitzii*. *A. communis* is linked to branched-chain amino acid metabolism and is enriched in people with metabolic disease^28^. We have previously found *Alistipes* species to be more prevalent in people with more severe dementia^11^, and others have found *Alistipes* species trigger inflammation^29^. *Guopingia tenuis* has been associated with resistance to diet-induced weight gain^30^ and may partially explain the lower body mass index observed in participants with higher Ap burden. *Bacteroides fragilis* and *Faecalibacterium prausnitzii* are commonly regarded as beneficial gut commensals^31^, with their presence in the cohort possibly indicative of the early and mild phase of disease. However, increased *Faecalibacterium prausnitzii* abundance has previously been reported in individuals with clinical AD^32^. Together, these findings indicate that enrichment of otherwise common or beneficial taxa may have context-dependent effects in early AD pathology.

Beyond compositional changes, we observed progressive reductions in predicted microbial metabolic capacity with increasing Aβ, particularly in relation to biosynthesis of SCFAs, amino acids, and neurotransmitters. SCFAs are microbially-produced immunomodulators and the main by-products from bacterial fermentation in the intestines, consisting mostly of acetate, butyrate, and propionate, and act locally on intestinal epithelial cells and immune cells to maintain gut barrier integrity and protect against inflammation^9^. Importantly, SCFAs can cross the blood–brain barrier and modulate microglial activation^33^, thereby limiting neuroinflammation and neurodegeneration. Reduced abundance of SCFA-producing bacteria has previously been associated with higher odds of AD pathology^34^ and more rapid cognitive decline^35^. Our findings extend this association to the preclinical stage, suggesting that impaired microbial immunomodulation may contribute to early Ap accumulation.

Earlier monitoring of AD pathology could enable identification of cognitively unimpaired individuals at elevated risk of progression, thereby extending the therapeutic window to the preclinical stage^16^. Currently, no disease-modifying interventions are approved for asymptomatic individuals^36^, as AD management remains centred on anti-amyloid monoclonal antibodies in early clinical disease^37^, pharmacological treatment of symptoms (e.g. cholinesterase inhibitors and memantine), behavioural and psychological symptom management (e.g. antipsychotics and sedatives), and non-pharmacological interventions, such as exercise. Population-based risk profiling using scalable biomarkers may substantially improve AD prevention and care by facilitating earlier intervention^36^. In this context, gut microbiome profiling has emerged as a promising non-invasive approach to assess brain health in older adults^38^. Stool-based screening is already embedded in public health programs for colorectal cancer prevention^39^, and microbiome-targeted interventions are increasingly feasible across diverse disease settings^40,41^. Growing evidence linking the microbiome–gut– brain axis to neurodegenerative diseases, including AD, further supports this approach, although investigations remain at an early stage^42,43^.

Despite these findings, it is important to acknowledge the cross-sectional design of this study as a limitation that precluded causal inference. While associations between the gut microbiome and dementia have been reported across disease stages^12–14^, AP deposition alone does not guarantee clinical progression^44^. Longitudinal studies spanning the AD continuum are therefore required to determine whether microbiome alterations contribute causally to Aβ accumulation and cognitive decline, and to evaluate the efficacy of microbiome-targeted interventions.

In conclusion, we report significant associations between increasing cerebral Aβ burden and gut microbiome composition and function, including pathways related to neurotransmission, immune regulation, and cellular metabolism, and in cognitively unimpaired individuals. These findings highlight the gut microbiome as a promising adjunct tool for AD risk stratification and early intervention.

## Data Availability

Data produced in the present study are available upon reasonable request to the authors.

## ACKNOWLEDGMENTS

We thank the participants and their study partners for their involvement and support of this research.

## CONSENT STATEMENT

All participants provided written informed consent. Approval for the study was obtained from the University of Adelaide Human Research Ethics Committee (H-2022-036). The study protocol was published on the Australian and New Zealand Clinical Trial Registry (ACTRN12622000891729).

