## Supplementary Material for "Change in gut microbiome precedes symptomatic Alzheimer’s disease and is correlated with amyloid-beta accumulation in the brain"

**Table S1:** **Alpha diversity of the gut microbiome of people undergoing testing for preclinical Alzheimer’s disease.** Unadjusted and multivariable-adjusted correlations between Aβ CL values and gut microbiome alpha diversity metrics (Shannon-Wiener diversity, Simpson’s index, Inverse Simpson’s index, Species richness). Multivariable analysis treated Aβ as a continuous variable and adjusted for age, sex, education, hypertension, diabetes, smoking, obesity, traumatic brain injury, depression, and high cholesterol.

|  | **Unadjusted  *p* value** | **Multivariable-adjusted  *p* value^a^** | **Multivariable-adjusted  *R* value** |
| --- | --- | --- | --- |
| **Shannon-Wiener diversity (*H’*)** | 0.758 | 0.913 | -0.012 |
| **Simpson’s index (*D*)** | 0.918 | 0.618 | -0.053 |
| **Inverse Simpson’s index (1/*D*)** | 0.918 | 0.618 | -0.053 |
| **Species richness (*d*)** | 0.559 | 0.823 | 0.024 |

^a^Age + sex + education + hypertension + diabetes + smoking + obesity + traumatic brain centre + depression + high cholesterol.

**Table S2:** **The** **gut microbial species significantly related to accumulated amyloid-beta in people undergoing testing for preclinical Alzheimer’s disease.** The magnitude of the difference in microbial species relative abundance with increasing accumulation of amyloid-beta presented as highest to lowest odds ratio with 95% confidence intervals following a multivariable adjustment. *Q* values report FDR-corrected statistical significance. Species with *q*<0.05 shown. Multivariable analysis treated amyloid-beta as a continuous variable and adjusted for age, sex, education, hypertension, diabetes, smoking, obesity, traumatic brain injury, depression, and high cholesterol. CI; confidence interval.

| **Species name** | **Odds Ratio  (95% CI)** | **Mean relative abundance  (%)** | ***Q* value** |
| --- | --- | --- | --- |
| *Anaerotignum faecicola* | 0.983  (0.978, 0.988) | 0.086 | <0.0001 |
| *Eubacterium rectale* | 0.989  (0.986, 0.993) | 2.024 | <0.0001 |
| *Clostridium SGB4909* | 0.990  (0.985, 0.994) | 0.033 | 0.0003 |
| *Streptococcus parasanguinis* | 0.990  (0.987, 0.994) | 0.085 | 0.0001 |
| *Phascolarctobacterium faecium* | 0.991  (0.985, 0.995) | 0.336 | 0.0026 |
| *Phocea massiliensis* | 0.991  (0.987, 0.995) | 0.005 | 0.0002 |
| *Actinomyces sp ICM47* | 0.991  (0.986, 0.996) | 0.003 | 0.0037 |
| *Mogibacterium diversum* | 0.991  (0.986, 0.996) | 0.003 | 0.0036 |
| *Ruminococcus bromii* | 0.991  (0.988, 0.995) | 2.529 | 0.0001 |
| *Anaerosacchariphilus sp NSJ 68* | 0.992  (0.988, 0.996) | 0.004 | 0.0012 |
| *Anaerobutyricum soehngenii* | 0.992  (0.989, 0.996) | 0.341 | 0.0003 |
| *Faecalicatena contorta* | 0.992  (0.988, 0.996) | 0.009 | 0.0020 |
| *Eubacterium ventriosum* | 0.992  (0.989, 0.996) | 0.148 | 0.0011 |
| *Enterocloster bolteae* | 0.993  (0.989, 0.997) | 0.053 | 0.0039 |
| *Bacterium 210917 DFI 7 65* | 0.993  (0.989, 0.997) | 0.011 | 0.0038 |
| *Candidatus Gastranaerophilales bacterium* | 0.993  (0.988, 0.997) | 0.146 | 0.0138 |
| *Roseburia sp AF02 12* | 0.994  (0.990, 0.997) | 0.250 | 0.0056 |
| *Hydrogenoanaerobacterium saccharovorans* | 0.994  (0.990, 0.997) | 0.026 | 0.0056 |
| *Blautia hydrogenotrophica* | 0.994  (0.989, 0.998) | 0.026 | 0.0158 |
| *Bacteroides thetaiotaomicron* | 0.994  (0.990, 0.997) | 0.508 | 0.0037 |
| *Lacrimispora amygdalina* | 0.994  (0.990, 0.997) | 0.081 | 0.0041 |
| *Vescimonas coprocola* | 0.994  (0.991, 0.997) | 0.165 | 0.0048 |
| *Lawsonibacter SGB15145* | 0.994  (0.991, 0.998) | 0.016 | 0.0056 |
| *Blautia obeum* | 0.994  (0.990, 0.998) | 0.523 | 0.0115 |
| *Oscillibacter sp ER4* | 0.994  (0.991, 0.998) | 0.520 | 0.0057 |
| *Dorea sp AF36 15AT* | 0.994  (0.991, 0.998) | 0.090 | 0.0129 |
| *Blautia wexlerae* | 0.995  (0.991, 0.998) | 1.215 | 0.0158 |
| *Butyricimonas paravirosa* | 0.995  (0.991, 0.998) | 0.043 | 0.0269 |
| *Streptococcus thermophilus* | 0.995  (0.991, 0.998) | 0.141 | 0.0269 |
| *Coprococcus catus* | 0.995  (0.991, 0.999) | 0.190 | 0.0291 |
| *Eggerthella lenta* | 0.995  (0.991, 0.998) | 0.047 | 0.0193 |
| *Dorea longicatena* | 0.995  (0.992, 0.999) | 0.865 | 0.0277 |
| *Alistipes putredinis* | 0.995  (0.991, 0.999) | 1.493 | 0.0348 |
| *Bacteroides caccae* | 0.995  (0.992, 0.999) | 0.568 | 0.0269 |
| *Coprococcus comes* | 0.995  (0.992, 0.999) | 0.456 | 0.0331 |
| *Flavonifractor plautii* | 0.995  (0.991, 0.999) | 0.081 | 0.0440 |
| *Clostridiaceae bacterium Marseille Q4145* | 0.995  (0.992, 0.999) | 0.025 | 0.0267 |
| *Roseburia inulinivorans* | 0.995  (0.992, 0.999) | 0.336 | 0.0269 |
| *Ruminococcus torques* | 0.996  (0.992, 0.999) | 0.733 | 0.0324 |
| *Blautia sp MCC283* | 1.005  (1.001, 1.008) | 0.254 | 0.0440 |
| *Methanobrevibacter smithii* | 1.005  (1.001, 1.008) | 0.570 | 0.0346 |
| *Lachnospira eligens* | 1.005  (1.001, 1.008) | 0.872 | 0.0370 |
| *Diplocloster agilis* | 1.005  (1.001, 1.009) | 0.001 | 0.0469 |
| *Blautia luti* | 1.005  (1.001, 1.009) | 0.062 | 0.0277 |
| *Butyricimonas faecihominis* | 1.005  (1.002, 1.009) | 0.051 | 0.0193 |
| *Cuneatibacter sp NSJ 177* | 1.006  (1.002, 1.009) | 0.005 | 0.0056 |
| *Alistipes senegalensis* | 1.006  (1.002, 1.010) | 0.041 | 0.0129 |
| *Faecalibacterium prausnitzii* | 1.006  (1.002, 1.010) | 3.032 | 0.0058 |
| *Lawsonibacter hominis* | 1.006  (1.002, 1.010) | 0.022 | 0.0132 |
| *Massilioclostridium coli* | 1.006  (1.003, 1.010) | 0.005 | 0.0056 |
| *Bifidobacterium longum* | 1.006  (1.003, 1.010) | 0.276 | 0.0056 |
| *Luoshenia tenuis* | 1.007  (1.003, 1.011) | 0.001 | 0.0056 |
| *Clostridium sp AF12 28* | 1.007  (1.003, 1.011) | 0.013 | 0.0060 |
| *Bacteroides fragilis* | 1.007  (1.003, 1.011) | 0.182 | 0.0020 |
| *Clostridium sp AF36 4* | 1.008  (1.004, 1.011) | 0.371 | 0.0003 |
| *Catenibacillus scindens* | 1.008  (1.004, 1.012) | 0.002 | 0.0010 |
| *Clostridiaceae bacterium Marseille Q4149* | 1.008  (1.005, 1.012) | 0.415 | 0.0002 |
| *Pseudoflavonifractor capillosus* | 1.009  (1.005, 1.013) | 0.010 | 0.0002 |
| *Blautia sp OF03 15BH* | 1.009  (1.005, 1.013) | 0.007 | 0.0001 |
| *Yeguia hominis* | 1.009  (1.006, 1.013) | 0.003 | <0.0001 |
| *Guopingia tenuis* | 1.010  (1.006, 1.014) | 0.009 | <0.0001 |
| *Alistipes communis* | 1.013  (1.009, 1.017) | 0.242 | <0.0001 |

**Table S3:** **Gut microbial metabolic capacity significantly related with accumulated amyloid-beta in people undergoing testing for preclinical Alzheimer’s disease.** The magnitude of the difference in microbial functional pathway relative abundance with increasing accumulation of amyloid-beta presented as highest to lowest odds ratio with 95% confidence intervals following a multivariable adjustment. *Q* values report FDR-corrected statistical significance. Pathways with *q*<0.05 shown. Multivariable analysis treated amyloid-beta as a continuous variable and adjusted for age, sex, education, hypertension, diabetes, smoking, obesity, traumatic brain injury, depression, and high cholesterol. CI; confidence interval.

| **Pathway (MetaCyc ID)** | **Function** | **Odds Ratio  (95% CI)** | **Mean relative abundance  (%)** | ***Q* value** |
| --- | --- | --- | --- | --- |
| PWY.6527 | Stachyose degradation | 0.987  (0.982, 0.991) | 0.000267 | <0.0001 |
| PWY.6317 | D-galactose degradation | 0.987  (0.983, 0.991) | 0.000297 | <0.0001 |
| COLANSYN.PWY | Colanic acid biosynthesis | 0.987  (0.983, 0.991) | 0.000148 | <0.0001 |
| HISTSYN.PWY | L-histidine biosynthesis | 0.989  (0.985, 0.993) | 0.000388 | <0.0001 |
| PANTO.PWY | Phosphopantothenate biosynthesis | 0.989  (0.985, 0.993) | 0.000327 | <0.0001 |
| THISYNARA.PWY | Thiamine diphosphate biosynthesis | 0.990  (0.986, 0.993) | 0.000172 | <0.0001 |
| PWY0.1061 | L-alanine biosynthesis | 0.990  (0.986, 0.994) | 0.000048 | 0.0001 |
| PWY0.1319 | CDP-diacylglycerol biosynthesis | 0.991  (0.987, 0.994) | 0.000425 | <0.0001 |
| PWY.5667 | CDP-diacylglycerol biosynthesis | 0.991  (0.987, 0.994) | 0.000425 | <0.0001 |
| PWY.6902 | Chitin degradation | 0.991  (0.987, 0.995) | 0.000074 | 0.0002 |
| FUCCAT.PWY | Fucose degradation | 0.991  (0.987, 0.995) | 0.000056 | 0.0002 |
| HSERMETANA.PWY | L-methionine biosynthesis | 0.991  (0.987, 0.995) | 0.000262 | 0.0004 |
| GLYCOLYSIS.I | Glycolysis | 0.992  (0.988, 0.995) | 0.000180 | 0.0003 |
| PWY.7883 | Anhydromuropeptides recycling | 0.992  (0.988, 0.995) | 0.000011 | 0.0003 |
| PWY.7456 | Mannan degradation | 0.992  (0.988, 0.995) | 0.000096 | 0.0002 |
| COMPLETE.ARO.PWY | Aromatic acid biosynthesis | 0.992  (0.988, 0.995) | 0.000489 | 0.0003 |
| PWY.7238 | Sucrose biosynthesis | 0.992  (0.988, 0.995) | 0.000602 | 0.0004 |
| PWY.7282 | Thiamine biosynthesis | 0.992  (0.988, 0.995) | 0.000164 | 0.0003 |
| COA.PWY | Coenzyme-A biosynthesis | 0.992  (0.988, 0.996) | 0.000403 | 0.0003 |
| FERMENTATION.PWY | Mixed acid fermentation | 0.992  (0.988, 0.996) | 0.000116 | 0.0006 |
| HEMESYN2.PWY | Heme biosynthesis | 0.992  (0.988, 0.996) | 0.000024 | 0.0007 |
| PWY.6629 | L-tryptophan biosynthesis | 0.992  (0.988, 0.996) | 0.000389 | 0.0006 |
| COA.PWY.1 | Coenzyme-A biosynthesis | 0.992  (0.988, 0.996) | 0.000423 | 0.0005 |
| PWY.724 | L-lysine, L-threonine, L-methionine biosynthesis | 0.992  (0.989, 0.996) | 0.000356 | 0.0007 |
| THISYN.PWY | Thiamine biosynthesis | 0.992  (0.989, 0.996) | 0.000218 | 0.0006 |
| PWY.5494 | Pyruvate fermentation | 0.993  (0.988, 0.997) | 0.000001 | 0.0031 |
| PROPFERM.PWY | L-alanine fermentation | 0.993  (0.988, 0.997) | 0.000002 | 0.0031 |
| PWY.8188 | L-alanine degradation | 0.993  (0.988, 0.997) | 0.000002 | 0.0031 |
| PWY.8189 | L-alanine degradation | 0.993  (0.988, 0.997) | 0.000002 | 0.0031 |
| PWY.5659 | GDP-mannose biosynthesis | 0.993  (0.989, 0.996) | 0.000130 | 0.0015 |
| PANTOSYN.PWY | Coenzyme-A biosynthesis | 0.993  (0.989, 0.996) | 0.000346 | 0.0011 |
| ANAEROFRUCAT.PWY | Homolactic fermentation | 0.993  (0.989, 0.996) | 0.000183 | 0.0019 |
| PWY.6163 | Chorismate biosynthesis | 0.993  (0.989, 0.997) | 0.000500 | 0.0020 |
| ARO.PWY | Chorismate biosynthesis | 0.993  (0.989, 0.996) | 0.000471 | 0.0015 |
| GLUTORN.PWY | L-ornithine biosynthesis | 0.993  (0.989, 0.997) | 0.000337 | 0.0015 |
| GLYCOGENSYNTH.PWY | Glycogen biosynthesis | 0.993  (0.989, 0.997) | 0.000520 | 0.0020 |
| P164.PWY | Purine nucleobases degradation | 0.993  (0.989, 0.997) | 0.000054 | 0.0019 |
| NONMEVIPP.PWY | Isopentenyl diphosphate biosynthesis | 0.993  (0.989, 0.997) | 0.000481 | 0.0019 |
| PWY.5484 | Glycolysis | 0.993  (0.989, 0.997) | 0.000168 | 0.0023 |
| GOLPDLCAT.PWY | Glycerol degradation | 0.993  (0.989, 0.997) | 0.000024 | 0.0019 |
| FUC.RHAMCAT.PWY | Fucose and rhamnose degradation | 0.993  (0.989, 0.997) | 0.000045 | 0.0019 |
| PWY.5941 | Glycogen degradation | 0.993  (0.989, 0.997) | 0.000435 | 0.0027 |
| PWY.7323 | O-antigen biosynthesis | 0.993  (0.990, 0.997) | 0.000106 | 0.0027 |
| PWY.7115 | Carbon dioxide fixation | 0.993  (0.989, 0.997) | 0.000038 | 0.0043 |
| PWY.6895 | Thiamine diphosphate biosynthesis | 0.993  (0.990, 0.997) | 0.000095 | 0.0031 |
| VALSYN.PWY | L-valine biosynthesis | 0.993  (0.990, 0.997) | 0.000511 | 0.0030 |
| PWY0.1277 | Acetyl coenzyme-A biosynthesis | 0.994  (0.989, 0.998) | 0.000010 | 0.0216 |
| PWY.3001 | L-isoleucine biosynthesis | 0.994  (0.990, 0.997) | 0.000297 | 0.0090 |
| PWY.6630 | L-tyrosine biosynthesis | 0.994  (0.990, 0.998) | 0.000105 | 0.0108 |
| RHAMCAT.PWY | L-rhamnose degradation | 0.994  (0.990, 0.997) | 0.000149 | 0.0056 |
| CALVIN.PWY | Calvin cycle | 0.994  (0.990, 0.997) | 0.000411 | 0.0034 |
| HCAMHPDEG.PWY | Propionate degradation | 0.994  (0.989, 0.998) | 0.000008 | 0.0225 |
| PWY.6690 | Phenolic compound degradation | 0.994  (0.989, 0.998) | 0.000008 | 0.0225 |
| THRESYN.PWY | L-threonine biosynthesis | 0.994  (0.990, 0.997) | 0.000291 | 0.0034 |
| PWY.7851 | Coenzyme-A biosynthesis | 0.994  (0.990, 0.997) | 0.000397 | 0.0050 |
| PWY.6470 | Peptidoglycan biosynthesis | 0.994  (0.990, 0.998) | 0.000079 | 0.0123 |
| POLYAMSYN.PWY | Polyamine biosynthesis | 0.994  (0.990, 0.998) | 0.000050 | 0.0139 |
| PWY0.1586 | Peptidoglycan biosynthesis | 0.994  (0.990, 0.998) | 0.000371 | 0.0130 |
| PWY.7400 | L-arginine biosynthesis | 0.994  (0.990, 0.998) | 0.000284 | 0.0135 |
| 1CMET2.PWY | Folate biosynthesis | 0.994  (0.991, 0.998) | 0.000385 | 0.0126 |
| PWY.621 | Sucrose degradation | 0.994  (0.990, 0.998) | 0.000134 | 0.0139 |
| PWY.6628 | L-phenylalanine biosynthesis | 0.994  (0.991, 0.998) | 0.000147 | 0.0142 |
| P41.PWY | Acetate and lactate biosynthesis | 0.994  (0.991, 0.998) | 0.000206 | 0.0139 |
| PWY.6121 | Purine nucleotide biosynthesis | 0.994  (0.990, 0.998) | 0.000438 | 0.0168 |
| PWY66.409 | Purine nucleotide biosynthesis | 0.994  (0.990, 0.998) | 0.000098 | 0.0204 |
| PWY.6823 | Molybdenum cofactor biosynthesis | 0.994  (0.991, 0.998) | 0.000165 | 0.0185 |
| ARGININE.SYN4.PWY | L-ornithine biosynthesis | 0.995  (0.991, 0.998) | 0.000128 | 0.0181 |
| PWY66.429 | Fatty acid biosynthesis | 0.995  (0.991, 0.998) | 0.000417 | 0.0132 |
| PYRIDNUCSYN.PWY | NAD biosynthesis | 0.995  (0.991, 0.998) | 0.000294 | 0.0176 |
| PWY.5384 | Sucrose degradation | 0.995  (0.991, 0.998) | 0.000132 | 0.0204 |
| ARG.POLYAMINE.SYN | Arginine and polyamine biosynthesis | 0.995  (0.991, 0.998) | 0.000080 | 0.0224 |
| PWY.5100 | Acetate and lactate biosynthesis | 0.995  (0.991, 0.998) | 0.000207 | 0.0204 |
| PWY.6892 | Thiamine diphosphate biosynthesis | 0.995  (0.991, 0.998) | 0.000209 | 0.0208 |
| PWY.822 | Fructan biosynthesis | 0.995  (0.991, 0.998) | 0.000002 | 0.0271 |
| PWY.7328 | O-antigen biosynthesis | 0.995  (0.991, 0.998) | 0.000138 | 0.0228 |
| GLCMANNANAUT.PWY | Amine and polyamine degradation | 0.995  (0.991, 0.998) | 0.000103 | 0.0213 |
| COBALSYN.PWY | Adenosylcobalamin biosynthesis | 0.995  (0.991, 0.998) | 0.000161 | 0.0224 |
| PWY.6531 | Mannitol cycle | 0.995  (0.991, 0.998) | 0.000021 | 0.0204 |
| PWY.5130 | 2-oxobutanoate degradation | 0.995  (0.991, 0.998) | 0.000025 | 0.0242 |
| PWY.5840 | Menaquinol biosynthesis | 0.995  (0.991, 0.998) | 0.000028 | 0.0219 |
| METH.ACETATE.PWY | Methanogenesis | 0.995  (0.991, 0.998) | 0.000004 | 0.0228 |
| PWY.6590 | Butanoate biosynthesis | 0.995  (0.991, 0.998) | 0.000039 | 0.0227 |
| HOMOSER.METSYN.PWY | L-methionine biosynthesis | 0.995  (0.991, 0.998) | 0.000021 | 0.0228 |
| MET.SAM.PWY | L-methionine biosynthesis | 0.995  (0.991, 0.998) | 0.000060 | 0.0228 |
| METSYN.PWY | L-homoserine and L-methionine biosynthesis | 0.995  (0.991, 0.998) | 0.000032 | 0.0228 |
| PWY.5347 | L-methionine biosynthesis | 0.995  (0.991, 0.998) | 0.000034 | 0.0228 |
| ANAGLYCOLYSIS.PWY | Glycolysis | 0.995  (0.991, 0.999) | 0.000452 | 0.0340 |
| GLUDEG.I.PWY | GABA degradation | 0.995  (0.991, 0.998) | 0.000031 | 0.0228 |
| PWY.2942 | L-lysine biosynthesis | 0.995  (0.991, 0.999) | 0.000357 | 0.0329 |
| PWY.6168 | Flavin biosynthesis | 0.995  (0.991, 0.999) | 0.000084 | 0.0268 |
| CENTFERM.PWY | Butanoate biosynthesis | 0.995  (0.991, 0.999) | 0.000031 | 0.0323 |
| P161.PWY | Acetate biosynthesis | 0.995  (0.991, 0.999) | 0.000026 | 0.0454 |
| PWY4LZ.257 | Acetate biosynthesis | 0.995  (0.991, 0.999) | 0.000027 | 0.0454 |
| GLUCOSE1PMETAB.PWY | Glucose degradation | 0.995  (0.992, 0.999) | 0.000131 | 0.0344 |
| RIBOSYN2.PWY | Flavin biosynthesis | 0.995  (0.992, 0.999) | 0.000285 | 0.0341 |
| PWY.7357 | Thiamine biosynthesis | 0.995  (0.992, 0.999) | 0.000277 | 0.0453 |
| PWY.6897 | Thiamine biosynthesis | 0.995  (0.992, 0.999) | 0.000228 | 0.0340 |
| P125.PWY | Butanediol biosynthesis | 0.995  (0.992, 0.999) | 0.000004 | 0.0442 |
| PWY.622 | Starch biosynthesis | 1.005  (1.001, 1.009) | 0.000016 | 0.0341 |
| PWY.5209 | Methanogenesis | 1.005  (1.001, 1.009) | 0.000004 | 0.0261 |
| PWY.7199 | Pyrimidine nucleotide biosynthesis | 1.005  (1.002, 1.009) | 0.000344 | 0.0224 |
| PWY.8187 | L-arginine degradation | 1.006  (1.002, 1.009) | 0.000111 | 0.0186 |
| PWY66.399 | Gluconeogenesis | 1.007  (1.003, 1.011) | 0.000097 | 0.0027 |
| FASYN.INITIAL.PWY | Fatty acid biosynthesis | 1.007  (1.003, 1.011) | 0.000002 | 0.0027 |
| PWY.4041 | Glutathione metabolism | 1.008  (1.004, 1.012) | 0.000082 | 0.0011 |
| PWY.5392 | Reductive TCA cycle | 1.008  (1.004, 1.012) | 0.000002 | 0.0013 |
| P124.PWY | Acetate and lactate biosynthesis | 1.008  (1.004, 1.012) | 0.000014 | 0.0005 |
| PWY.7761 | NAD salvage pathway | 1.009  (1.005, 1.013) | 0.000113 | 0.0001 |
